# The Antiviral Mechanism of Action of Molnupiravir in Humans with COVID-19

**DOI:** 10.1101/2023.11.21.23298766

**Authors:** Timothy P. Sheahan, Laura J. Stevens, Tara M. Narowski, Taylor J. Krajewski, Chanhwa Lee, Katie R. Mollan, Jennifer Gribble, Fernando R. Moreira, Izabella N. Castillo, Edwing Cuadra, Paul Alabanza, Amy James Loftis, Robert W. Coombs, Erin A. Goecker, Alexander L. Greninger, James D. Chappell, Ariane J. Brown, John Won, Felicia Lipansky, Wayne Holman, Laura J. Szewczyk, Ralph S. Baric, Wendy P. Painter, Joseph J. Eron, Lakshmanane Premkumar, Mark R. Denison, William A. Fischer

**Affiliations:** Department of Epidemiology, Gillings School of Global Public Health, University of North Carolina at Chapel Hill, Chapel Hill, NC, USA; Department of Microbiology and Immunology, University of North Carolina at Chapel Hill, Chapel Hill, NC, USA; Department of Pediatrics, Vanderbilt University Medical Center, Nashville, TN, USA; Department of Biostatistics, Gillings School of Global Public Health, University of North Carolina at Chapel Hill, Chapel Hill, NC, USA; Institute for Global Health and Infectious Diseases, Division of Pulmonary Diseases and Critical Care Medicine, University of North Carolina at Chapel Hill, Chapel Hill, NC, USA; Department of Laboratory Medicine and Pathology, University of Washington, Seattle, WA, USA; Ridgeback Biotherapeutics LP, Miami, FL, USA; Department of Medicine, Division of Infectious Diseases, University of North Carolina at Chapel Hill, Chapel Hill, NC, USA

**Author notes:** These authors contributed equally (Timothy P. Sheahan, William A. Fischer). Co-corresponding authors: WAF, TPS.

## Abstract

Meaningful metrics of antiviral activity are essential for determining the efficacy of therapeutics in human clinical trials. Molnupiravir (MOV) is a broadly acting antiviral nucleoside analog prodrug that acts as a competitive alternative substrate for the SARS-CoV-2 RNA-dependent RNA polymerase (RdRp). We developed an assay, Culture-PCR, to better understand the impact of MOV therapy on infectious SARS-CoV-2. Culture-PCR revealed MOV eliminated infectious virus within 48 hours in the nasopharyngeal compartment, the upper airway location with the greatest levels of infectious virus. MOV therapy was associated with increases in mutations across the viral genome but select regions were completely unaffected, thus identifying regions where mutation likely abrogates infectivity. MOV therapy did not alter the magnitude or neutralization capacity of the humoral immune response, a documented correlate of protection. Thus, we provide holistic insights into the function of MOV in adults with COVID-19.

## Introduction

Severe acute respiratory syndrome coronavirus 2 (SARS-CoV-2), the etiologic agent of coronavirus disease 2019 (COVID-19), has caused more than 771 million infections and over 6.9 million deaths worldwide as of October, 2023. Several SARS-CoV-2 vaccines are available yet despite both infection-or vaccine-induced immunity, the persistent circulation and evolution of SARS-CoV-2 variants of concern (VOC) have caused frequent breakthrough infections. Waves of VOC evolution in the spike protein have facilitated escape from all previously authorized therapeutic monoclonal antibodies to date, rendering them clinically ineffective. Recent reports suggest that mAb therapies could also interfere with the development of an endogenous SARS-CoV-2-specific humoral immune response^1–3^. Approved direct-acting antiviral therapies also have treatment challenges, including the intravenously administered remdesivir and complex drug-drug interactions associated with nirmatrelvir/ritonavir and ensitrelvir^4^. Additionally, there is a continued need for safe and effective oral antivirals and to understand their impact on adaptive immunity.

Robust and biologically relevant measures of antiviral activity in humans are critical to understand the effectiveness of antiviral therapies in clinical trials and the potential impact of viral genomic changes. Molnupiravir (MOV) is a prodrug of N-hydroxycytidine with broad antiviral activity against acute SARS-CoV-2 and related VOC, SARS-CoV, MERS-CoV, and several other human and enzootic CoV infections *in vitro* and *in vivo*^5,6^. MOV also attenuates SARS-CoV-2 infection associated chronic organizing pneumonia with fibrotic lesions in mice^7^.

Based on in vitro and animal studies, MOV is believed to exert its antiviral activity by inducing random mutations in viral genomic and subgenomic RNA, thus driving virus populations to extinction ^6–11^. One study has proposed that MOV therapy was associated with increased mutations in SARS-CoV-2 RNA, but this analysis did not investigate how infectious virus populations were affected or how a rapid reduction in viral replication affects the development of a protective humoral immune response ^6^. More recently, a second bioinformatic study analyzing global SARS-CoV-2 sequence data argued that MOV use may be associated with the transmission of viruses containing a MOV-associated mutational signature without direct clinical data to demonstrate sequences arose in patients on MOV treatment, epidemiologic data to plausibly link the patients in which sequences were derived or virologic data to determine if sequences were associated with infectious virus^12^.

Most clinical trials of antiviral therapies for COVID-19 rely on the measurement of viral RNA via reverse transcription polymerase chain reaction (RT-PCR) to determine antiviral activity, but with this assay, it is not possible to determine if the measured viral RNA is associated with infectious virus particles, defective virus particles, antibody neutralized virus particles or sloughed infected cells. To best evaluate the antiviral activity of MOV on infectious virus, we developed a virus culture/RT-PCR (Culture-PCR) assay to sensitively detect infectious SARS-CoV-2 in respiratory samples from a randomized placebo-controlled Phase 2a trial in outpatients with mild-to-moderate COVID-19 conducted June 2020 through January 2021, prior to widespread vaccination and the emergence of the alpha VOC ^9^. Using Culture-PCR, we determined that the nasopharyngeal compartment had the greatest levels of SARS-CoV-2 infectious virus and that MOV exerted a dose-dependent antiviral effect. Deep sequencing of infectious virus populations from those randomized to Placebo and MOV revealed that MOV therapy was associated with increases in mutations across the viral genome but select regions were completely unaffected, thus identifying regions where mutation likely abrogates infectivity. Here, we build upon our prior work to better understand the compartmentalization of SARS-CoV-2 replication in the upper airway, the MOV mechanism of action *in vivo*, and the potential impact of therapy on the humoral immune response.

## Results

### Culture-PCR reveals compartmentalization of infectious SARS-CoV-2 in the upper airway

We aimed to comprehensively characterize the antiviral activity and mechanism of action (MOA) of MOV in humans. To achieve this goal, we needed to first develop assays to detect infectious SARS-CoV-2 in human respiratory samples and then determine the optimal respiratory compartment for the sampling of infectious virus. We developed two assays within which to assess the presence of infectious SARS-CoV-2 in human respiratory samples: visualizing cytopathic effect (CPE) via light microscopy and Culture-PCR. Using recombinant control SARS-CoV-2 (Supplementary Figure 1A-E), Culture-PCR was a more sensitive measure of infectious virus isolation over the monitoring of CPE. To identify the upper airway compartment with the greatest levels of infectious SARS-CoV-2, we measured viral RNA and infectivity in paired anterior nasal (AN), nasopharyngeal (NP) and oropharyngeal (OP) swabs (Figure 1A) collected from participants treated with Placebo or 200mg MOV in the Phase 2a study noted above (Figure 1B) ^9^. On visit day 1, levels of viral RNA in NP swabs were higher than that in AN (p = 0.0007) and OP swabs (p = 0.0002), and viral RNA levels tended to be higher in AN swab compared to OP swab (p=0.0593). Similar to our previous studies, viral RNA levels were predictive of Culture-PCR positivity in samples from all three compartments (NP, p = < 0.0001; AN, p = 0.0002; OP p = 0.0007) (Figure 1C) ^13^. NP swabs had the greatest proportion of infectious virus positive samples on all visit days assessed (Figure1D). Thus, the NP compartment offered the most sensitive means of detecting infectious virus.

**Figure 1.**
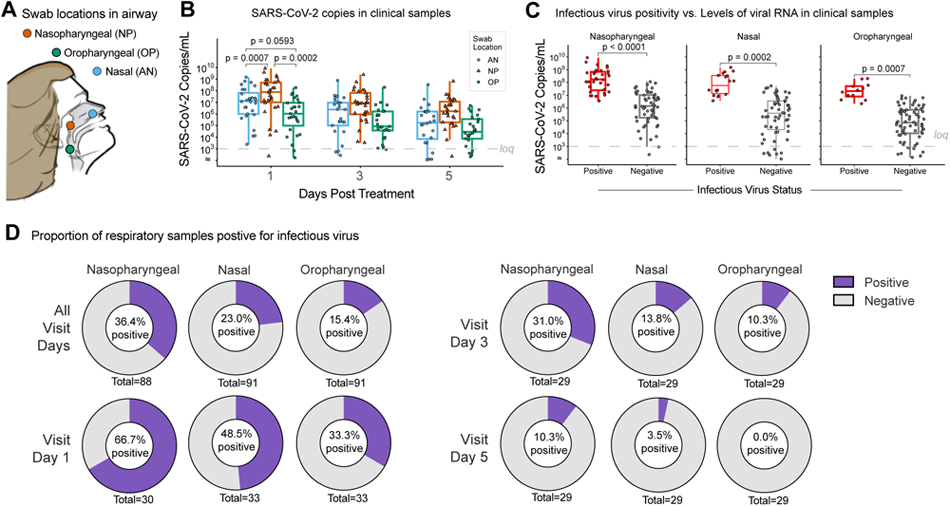
Culture-PCR reveals a compartmentalization of infectious SARS-CoV-2 in the upper airway. **(A)** Schematic showing the anatomical sites for respiratory sample collection. Nasal (AN), Nasopharyngeal (NP), and Oropharyngeal (OP). **(B)** SARS-CoV-2 viral RNA copies/ml in clinical samples collected from a subset of Placebo and 200mg MOV treated participants on visit days 1, 3, and 5 for each swab location. Sample size varies by swab location and day: day 1 (AN: n=28, NP: n=30, OP: n=27); day 3 (AN: n=25, NP: n=29, OP: n=25); day 5 (AN: n=24, NP: n=29, OP: n=25). SARS-CoV-2 RNA levels were compared using Wilcoxon signed-rank tests and P-values are displayed. **(C)** Infectious virus positivity compared to the levels of SARS-CoV-2 viral RNA in clinical samples from days 1, 3, and 5 combined for each swab location. Sample size varies by swab location and infectious positivity: NP (Positive: n=32, Negative: n=56); AN (Positive: n=18, Negative: n=59); OP (Positive: n=12, Negative: n=65). Wilcoxon rank-sum test P-value for clustered data (Datta-Satten method) to account for repeated measures correlation across the 3 days are displayed. For **B** and **C**, the boxes encompass 25th-75th percentile, the line is at the median and the whiskers extend to the extrema (no more than 1.5 times the IQR from the box). The RT-PCR assay limit of quantification (loq, 10^3^ Copies/mL) is represented by the grey dashed line; results <loq were handled as tied values for statistical analysis and for data visualization were randomly jittered below the loq. **(D)** The proportion of samples positive for infectious virus per swab site per visit day.

To confirm that Culture-PCR was superior to the visualization of CPE to assess infectivity, we screened over 500 NP swab samples from the MOV Phase 2A clinical trial noted above using these two methods (Fig. 2A). We observed a MOV dose-dependent effect on sample infectivity by both methods (Fig. 2B-E). In samples from those randomized to Placebo (Fig. 2B), the proportion of samples positive for infectious virus decreased over time (% Positive: Day 1 = 47.2 %; Day 3 = 17.0 %, Day 5 = 11.1 %) likely due to the induction of successful host immune responses. In addition, the proportion of infectious virus positive samples was greater via Culture-PCR than observation of CPE at all timepoints (Fig. 2B). MOV therapy at the FDA authorized dose of 800mg rapidly diminished infectious virus in NP swab samples after only 48hr of treatment with only 1 of 51 samples testing positive on visit day 3 and 0 of 51 samples testing positive on visit day 5 (Fig. 2C). Prior analysis demonstrated that MOV therapy with 800mg diminished infectious virus more rapidly than those randomized to Placebo on both visit day 3 and 5^9^. Similar trends were observed in those randomized to 400mg MOV (Fig. 2D), although statistical significance in infectious virus reduction was delayed as compared to those on 800mg and only observed on visit day 5^9^. The 200mg dose was found to be subtherapeutic and trends in infectivity were not different than those randomized to Placebo (Fig. 2E).

**Figure 2.**
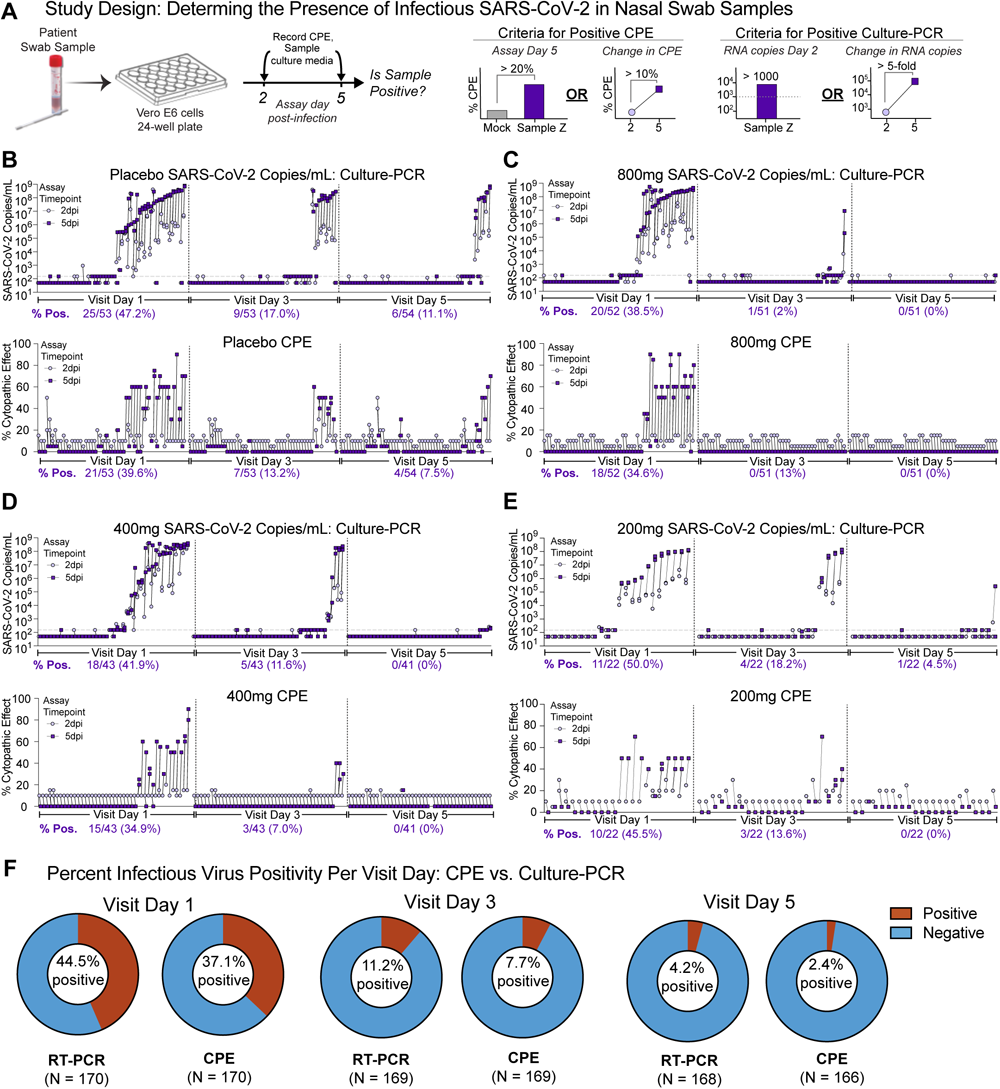
Culture PCR is a more sensitive means of determining infectivity in human respiratory samples than monitoring CPE. **(A)** Study design schematic and criteria for positivity. Culture PCR data (top) and CPE data for the Placebo **(B)**, 800mg **(C)**, 400mg **(D)** and 200mg **(E)** participants at baseline, visit Day 3 and 5. **(F)** The proportion of samples positive for infectious virus by Culture PCR positive or CPE for visit Days 1, 3 and 5. Includes data from Placebo, 200, 400 and 800mg participants.

Importantly, in all dose groups, Culture-PCR identified infectious SARS-CoV-2 in a greater proportion of samples at all times assessed (Fig. 2F). All together, these data indicate that Culture-PCR is a sensitive means of detecting infectious SARS-CoV-2, the NP compartment is the upper respiratory site with the greatest levels of infectious virus and that MOV therapy rapidly reduces infectious virus in the airway within 48hr of initiating treatment.

### MOV therapy is associated with an increase in SARS-CoV-2 RNA mutation frequency in infectious virus

MOV is a ribonucleoside analog antiviral which when metabolized to N4-hydroxycytidine 5’-triphosphate competes with intracellular cytidine 5’-triphosphate for inclusion in nascent viral RNA during virus replication. When incorporated into the template RNA strand it can be read as either a cytidine or uridine leading to an accumulation of transition mutations ^6^. To better understand the antiviral MOA in humans, we evaluated the impact of MOV therapy on SARS-CoV-2 genomic RNA isolated from infectious virus populations. We identified participants randomized to Placebo (n = 9) and MOV (200mg, n = 4; 400mg, n = 4; 800mg, n = 1) treatment groups with infectious virus positive respiratory samples by Culture-PCR on at least two timepoints including baseline and either day 3 or day 5 to determine the effect of treatment and time on SARS-CoV-2 viral RNA. As noted above, several participants in the Placebo group were positive for infectious virus on day 3 (9 of 54) and beyond, but few of those in the dose groups (400 and 800mg) proven to be therapeutically effective were, thus limiting the samples available for this analysis^9^. Infectious virus populations were isolated on Vero cells and total RNA from infected cultures was deep sequenced (Figure 3A). The resultant sequencing data was compared to SARS-CoV-2 WA1 to determine the numbers of nonsynonymous, synonymous, transition and transversion mutations in each virus population per patient per time point using 0.1% as our cutoff for positivity (Supplementary Figure 2). At baseline (day 1), prior to the initiation of MOV therapy or Placebo, similar numbers of mutations were observed in infectious viruses from participants randomized to receive Placebo or MOV (Figure 3B). In virus populations isolated from samples on visit day 3, we observed significant increases in nonsynonymous, synonymous, and transition mutations in participants who received MOV compared with Placebo (Figure 3B). G:A transition mutations were enriched in viruses from MOV-treated participants (Supplementary Figure 3) ^14^.

**Figure 3.**
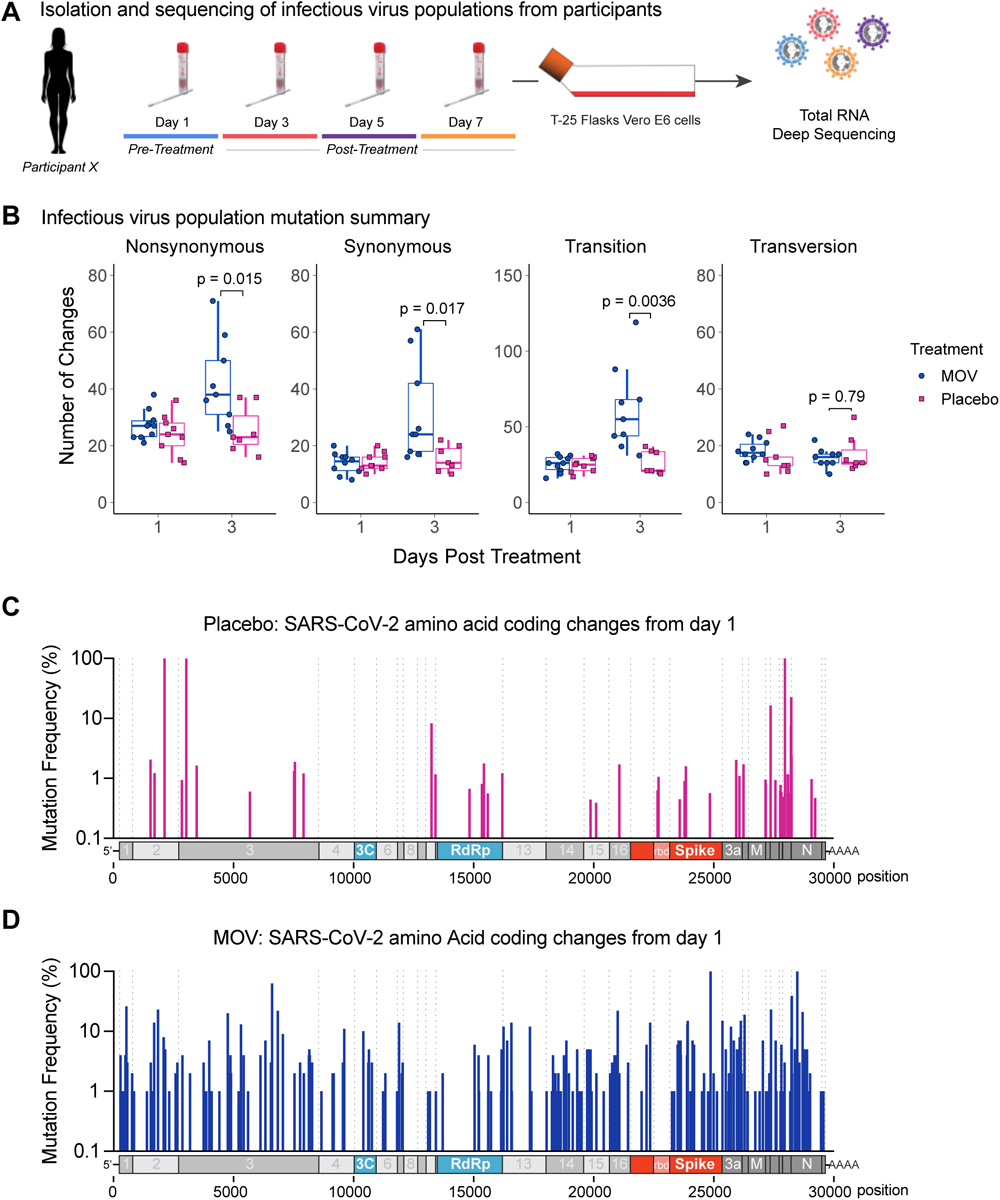
MOV therapy is associated with an increase in SARS-CoV-2 RNA mutation frequency in recoverable infectious virus. **(A)** Schematic for infectious virus isolation and deep sequencing. **(B)** SARS-CoV-2 infectious virus population mutation summary. Nonsynonymous, synonymous, transition, and transversion mutations are displayed by treatment group (MOV and Placebo); each dot represents a participant. The boxes encompass 25th-75th percentile, the line is at the median and the whiskers extend to the extrema (no more than 1.5 times the IQR from the box). For visit day 3, the number of changes were compared using Wilcoxon rank-sum tests and P-values are displayed. Sample size varies by visit day and treatment group. Day 1 (MOV: n=9, Placebo: n=9); day 3 (MOV: n=9, Placebo: n=7). **(C)** Placebo group SARS-CoV-2 amino acid coding changes unique to Day 3. Data for all 7 patients with infectious virus on day 3 are displayed. **(D)** MOV group SARS-CoV-2 coding changes unique to Day 3. Data for all 9 patients with infectious virus on day 3 are displayed. For **C** and **D**, the location and frequency of coding changes in infectious virus populations isolated from day 3 NP swab samples that differed from those observed in virus populations on day 1.

Since nonsynonymous coding changes have the potential to impact protein function, we mapped all nonsynonymous coding changes onto the SARS-CoV-2 genome. At baseline, coding changes were found throughout the genome in infectious virus populations from both Placebo and MOV-treated participants (Supplementary Figure 4). Coding changes were more common in structural and accessory open reading frame (ORF) genes located in the 3’ end of the genome compared to the 5’ ORF1a/b, which encodes several nonstructural proteins (Nsps) essential for replication and RNA synthesis. We then mapped the coding changes different than those observed on day 1 onto the SARS-CoV-2 genome to understand if there were regions of the genome that were tolerant and intolerant of change in infectious SARS-CoV-2 (Supplementary Figure 5). Coding changes on day 3 unique from day 1 were observed in Placebo group virus populations and like those at baseline prior to the initiation of therapy, tended to be greater in the 3’ end of the genome vs. ORF1a/b (Supplementary Fig. 5). Similarly, viruses from MOV-treated participants had a greater number of coding changes in the 3’end, however, the numbers of coding changes observed was increased compared to Placebo treated participants (Supplementary Fig. 5). Very few coding changes in the spike receptor binding domain (RBD) were observed in infectious viruses from both Placebo and MOV-treated participants (Supplementary Fig. 6). To better understand the plasticity of the infectious SARS-CoV-2 genome in Placebo and MOV treated participants, we mapped aggregated non-synonymous changes unique to day 3 on the viral genome (Fig. 3C and D). The small number of unique coding changes in viruses from Placebo on day 3 limited analysis of regions that were relatively spared from change, although a cluster of changes in ORFs 7a/b and ORF8 was noted suggesting these regions can change while retaining infectivity (Fig. 3C). In virus populations isolated from participants who received MOV, there was heterogeneity in the location of mutations. Numerous changes were observed in the aggregated data from all MOV-treated participants (Fig. 3D). For example, Nsp1, Nsp14-16, the C-terminus of Spike (S), 3a, ORF6, 7a/b, 8 and Nucleocapsid (N) all harbored multiple changes. However, the N-terminus of Nsp2, Nsp8, Nsp9, most of Nsp12 (RNA dependent RNA polymerase, RdRp), most of helicase (Nsp13), the S1 segment of S, and the C-terminus of N had few to no changes (Fig. 3D). Collectively, these data indicate that regions of the SARS-CoV-2 genome intolerant of change are likely essential to the retaining of infectivity.

### Humoral responses are unaffected by effective MOV therapy

Successful antiviral therapy is associated with an elimination of infectious virus as we observed in our Phase 2a trial in participants who received either 400 or 800mg MOV^9^. The reduction in viral protein antigens from antiviral therapy theoretically has the potential to negatively impact the humoral immune response and increase the risk of SARS-CoV-2 reinfection. To determine if MOV therapy at 400 or 800mg affected the humoral response, we measured the levels of IgM, IgA, and IgG targeting SARS-CoV-2 spike and nucleocapsid in plasma samples prior to the initiation of therapy (i.e. Day 1), and in those collected Days 7, 14 and 28 (Fig. 4). At baseline, differences in levels of SARS-CoV-2 RBD IgM, IgA, IgG or N-specific IgG between participants randomized to receive Placebo, MOV 400mg or MOV 800mg were not observed. IgA and IgM levels increased on days 7 and 14 and were statistically greater in participants who received Placebo compared to participants who received MOV 800mg but this difference disappeared by day 14. As expected, we observed increases in SARS-CoV-2 RBD and N-specific IgG over time. Importantly, we did not observe quantitative differences in antigen-specific antibody levels among MOV-treated groups (400 and 800mg) and Placebo (Fig. 4). We then used an ACE2 blockade assay, to quantify the levels and breadth of neutralizing antibodies in plasma. Importantly, 400 or 800mg of MOV therapy was comparable to the Placebo group for the generation of ACE2 receptor inhibiting antibodies against Wuhan, Alpha, Beta and Delta SARS-CoV-2 strains (Fig. 5). Neutralizing antibodies against more contemporary Omicron VOC subvariant lineages were largely absent in both the treatment and Placebo group, which is consistent with other studies conducted prior to the emergence of Omicron (Fig. 5). Collectively, these data demonstrate the kinetics, magnitude, and breadth of the humoral response were not impacted by successful MOV therapy.

**Figure 4.**
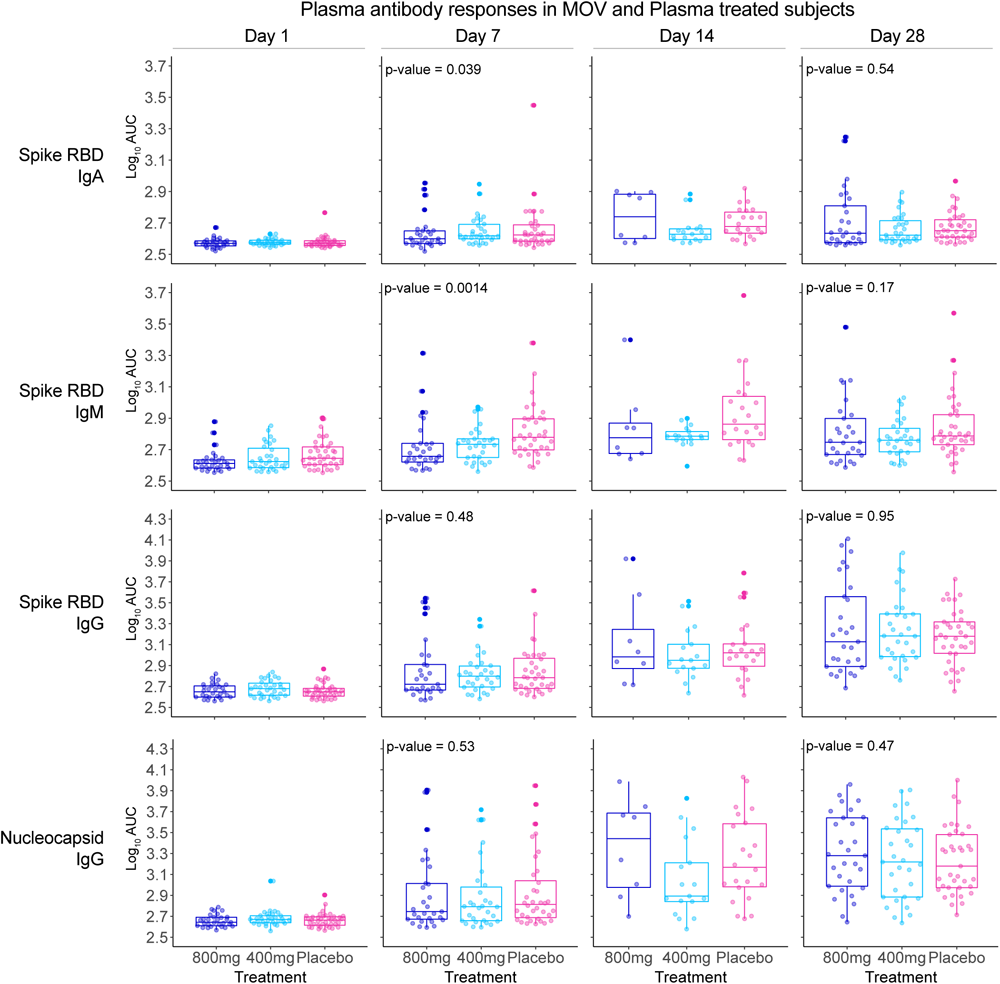
Plasma antibody responses are similar with MOV therapy. Antibody titer levels on visit days 1, 7, 14, and 28 for Spike RBD IgA, Spike RBD IgM, Spike RBD IgG and Nucleocapsid IgG are displayed by treatment dose level for participants seronegative by RBD IgG on day 1. Each dot represents a participant. The boxes encompass 25th-75th percentile, the line is at the median and the whiskers extend to the extrema (no more than 1.5 times the IQR from the box). P-values corresponding to Jonckheere-Terpstra (JT) test for dose level trend are displayed for day 7 and day 28. Sample size varies by dose group and visit day. Day 1 (800mg n=29, 400mg n=31, Placebo n=38); day 7 (800mg n=29, 400mg n=30, Placebo n=36); day 14 (800mg n=8, 400mg n=17, Placebo n=22); day 28 (800mg n=29, 400mg n=31, Placebo n=37).

**Figure 5.**
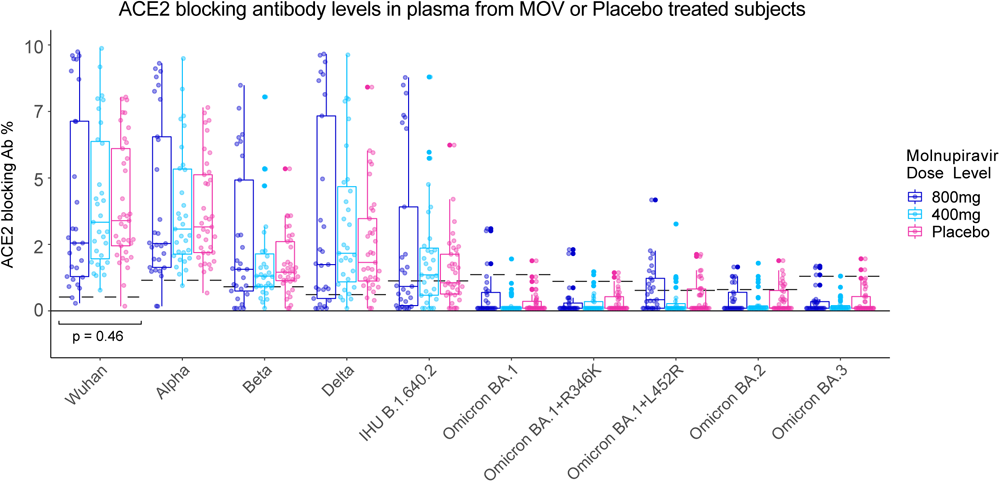
ACE2 blocking antibody levels are similar across dose groups. ACE2 Antibody (Ab) blocking % for each strain on visit day 28 displayed by treatment dose level for participants seronegative by RBD IgG on day 1; each dot represents a participant. P-values corresponding to Jonckheere-Terpstra (JT) test for dose level trend are displayed for Wuhan strain. Dashed lines display assay cut-offs for positivity. All ACE2 blocking Ab % values below 1% were set to 1%.

## Discussion

Since the emergence of SARS-CoV-2 in 2019, as of October 2023, there have been over 771 million infections and over 6.9 million deaths worldwide. Insufficient vaccine uptake, waning vaccine and infection-derived immunity, and the persistent circulation of SARS-CoV-2 have enabled the constant evolution and emergence of several variants that evade the protective efficacy of mAb therapeutics. Thus, potent, broadly-acting oral antivirals targeting the CoV family are needed to treat those at high risk for severe COVID-19 as well as future emerging CoV infections. For an antiviral like MOV, whose mechanism of action is driven by error induction in viral RNA replication, the true antiviral effect of the drug may be masked by only measuring changes in viral RNA by RT-PCR alone and thus requires the evaluation of the impact on infectious virus populations. Here we show that i) Culture-PCR is a more sensitive means of detecting infectious virus compared to detection of cytopathic effects by microscopy; ii) the nasopharynx has the highest levels of SARS-CoV-2 replication in the upper respiratory tract; iii) successful MOV therapy is associated with reductions in infectious virus in the nasopharynx and increased frequencies of nonsynonymous, synonymous, and transition mutations in the infectious viral genome; iv) by mapping the mutations from infectious virus isolated from MOV-treated participants, we identified regions in SARS-CoV-2 that may and may not tolerate change, providing insight into the evolutionary flexibility of genes, residues and proteins; v) despite a robust antiviral effect in all compartments by MOV, there appears to be no impairment in the development of a humoral IgG immune response.

Earlier *in vitro* and animal studies evaluating the antiviral activity of MOV against MERS-CoV found that MOV induces viral replication errors driven by an accumulation of transition mutations and recent biochemical data demonstrated that this primarily occurs in the template strand^6,15^. CoV encode a viral enzyme with proofreading activity, nsp14-ExoN, which mediates CoV replication fidelity and limits mutagenesis by multiple RNA mutagens including ribavirin and 5-fluorouracil *in vitro*^16^. In contrast, MOV can overcome or bypass nsp14-ExoN proofreading, driving increased mutation frequency in the SARS-CoV genome during replication^5^. This MOV MOA has similarly been observed for alphavirus and influenza virus in cell culture and in animal models ^17,18^. Here, we provide data from infectious virus populations from humans treated with MOV demonstrating an increase in transition mutations, resulting in both nonsynonymous and synonymous changes compared with Placebo-treated participants. To understand the impact of MOV on SARS-CoV-2 RNA species and the potential selection for drug-resistant viruses or novel VOC, the sample utilized for sequencing and the associated context/metadata are critically important. Recently, Donovan-Banefield et al. described the mutational spectra in total RNA from NP swabs from participants on MOV therapy from the AGILE trial in the United Kingdom which ran from November 2020 to March 2022^19^. One important differentiating factor from our work is the sample utilized for sequencing – the AGILE trial used total RNA from NP swabs while we evaluated RNA from infectious virus populations. In Donovan-Banefield et al., increases in G:A and C:U transitions were reported without enrichment of mutations within specific amino acids. In contrast, we found an increase in G:A mutations in specific regions in infectious virus. In addition, we found that specific regions in the infectious viral genome were tolerant or intolerant of mutational change. The differences in input sample may account for the differences observed in our collective patterns of transition mutations as compared to those reported by the AGILE trial. Taken together, the data presented here supports viral error induction as one *in vivo* mechanism of action. There are also recent reports of increases in transition mutations, like those observed in MOV-treated patients, in SARS-CoV-2 sequence databases (e.g. GISAID) and these increases were coincident with the widespread use of MOV in certain locales^20^. Although one can speculate about the origin of these mutations, without more information and/or epidemiologic investigations, it is not possible to determine if these sequences were derived from humans on MOV, whether viral RNA sequenced was from infectious or non-infectious viral particles/RNAs or if similar genomes purportedly associated with transmission events were epidemiologically related.

With the continued evolution of SARS-CoV-2 VOC over the course of the pandemic, this virus has already demonstrated a surprising capacity for change, continuously finding evolutionary paths which can evade neutralization by monoclonal antibody therapy and survive in populations with preexisting immunity. In addition, with the increased use of antiviral monotherapy, there is potential for the development of antiviral resistance. Thus, understanding capacity for change in infectious SARS-CoV-2 is important for rational drug development and screening for drug-resistance. Our current study and the prior Phase 2a study demonstrated that MOV therapy exerts a dose-dependent robust antiviral effect on viral RNA and infectious virus in the nasopharynx. For these reasons, we could only recover infectious virus from a single participant (1 of 53) treated with the most effective therapeutic dose (i.e. 800mg, the approved dose) on day 3, 48hr after treatment initiation and no infectious virus was detected in any participant treated with 800mg (0 of 53) on day 5. However, infectious virus was recoverable from 4 of 22 participants administered the subtherapeutic dose of 200mg on day 3 and 1 of 22 participants on day 5. We leveraged the subtherapeutic dosing groups to investigate the evolutionary plasticity of SARS-CoV-2 and show that certain relatively conserved genes coding for nonstructural proteins (e.g. Nsp1, 2, 3, 14, 16, etc.) demonstrate increased mutational frequency among infectious virus, while others had no detected mutations, suggesting that some regions may be more genetically flexible (e.g. Nsp8, 9, 12, 13). In addition, like the findings of Donovan-Banefield et al., we found an enrichment of mutations in the 3’ end of the genomes of infectious virus populations. Importantly, MOV treatment did not select for changes in the RdRp at or near locations of known nucleoside antiviral resistance or in the S RBD. These findings in combination with the rapid elimination of infectious virus after only 48 hours of treatment make it highly unlikely that MOV treatment could generate a VOC capable of widespread transmission.

Despite a robust antiviral effect, MOV treatment does not affect the magnitude and breadth of the SARS-CoV-2 humoral immune response, which are associated with protection against repeat infections and severe COVID-19. Prior studies of monoclonal antibody therapies demonstrated a treatment-associated suppression of the endogenous anti-SARSCoV-2 spike IgM response by 85-90% ^3^. The rapid reduction in viral load in the early stage of infection by mAb therapies likely plays a role in the reduced endogenous response. A decrease in spike IgM levels was seen on day 7 in MOV treated individuals compared with Placebo recipients, but this difference was transient and disappeared on Day 14 and 28. Unlike other antivirals like nirmatrelvir (i.e. Paxlovid), a viral protease inhibitor, or remdesivir (i.e. Velkury), a chain terminating nucleoside analog, the disparate MOA of MOV (i.e. viral error induction) may provide a benefit to the humoral immune response by facilitating the prolonged translation of impaired protein antigens from non-infectious genomes driving a relatively normal antibody response ^9^. The mechanism of action of MOV is thus likely the result of both preservation of a protective endogenous humoral immune response and rapid reduction in infectious virus populations.

Broad-spectrum antivirals are needed to treat people at high risk of developing severe COVID-19 and immunocompromised individuals that may experience prolonged SARS-CoV-2 replication. To robustly evaluate antiviral drug activity in humans, a thorough evaluation of meaningful metrics of antiviral activity, mechanism of action and potential immune interference are needed. Here, we provide a strategy using MOV as an example that could be applied to other small molecules and monoclonal antibodies. With the inevitable rise of new SARS-CoV-2 VOC, it is important to directly analyze patient samples, particularly for replication competent virus to allow direct understanding of the mechanisms and outcome of antiviral treatment.

## Data Availability

All data produced in the present study are available upon reasonable request to the authors

## Acknowledgements

The authors wish to acknowledge the participants and the following sites for their dedication and contributions to the study: Valley Clinical Trials (Northridge, CA), University of North Carolina (Chapel Hill, NC), Fred Hutchinson (Seattle, WA), Care United Research (Forney, TX), Benchmark Research (Colton, CA), FOMAT Medical Research (Oxnard, CA), Indago Research & Health Center (Hiahleah, FL), Wake Forest University (Winston Salem, NC), Duke University (Durham, NC), and NOLA Research Works (New Orleans, LA). Additionally, the authors acknowledge the randomization support provided by the National Center for Advancing Translational Sciences (NCATS; UL1TR002489) and the UNC Center for AIDS Research (P30 AI050410). Molnupiravir was invented at Drug Innovations at Emory (DRIVE) LLC, a not for profit biotechnology company wholly owned by Emory University, and with partial funding support from the US government.

## Materials and Methods

### Clinical Samples and Study Design

The clinical samples utilized in our study were obtained from a previously described Phase 2a randomized, double-blind, placebo-controlled study of multiple doses of molnupiravir in non-hospitalized adults with recently diagnosed COVID-19^9^. The patient population was comprised of unvaccinated adults 18 years of age and older with mild to moderate COVID-19 symptom onset within 7 days and who were PCR positive for SARS-CoV-2 infection within 96 hours at the time of enrollment (day 1) ^13^. We evaluated nasopharyngeal (NP) swabs collected on days 1 (baseline), 3 and 5 from 175 participants. For a subset of patients (N = 33), nasal (AN) and oropharyngeal (OP) swabs were concurrently collected with NP swabs. Nasal swabs collected from each participant at enrollment were placed into virus transport medium (VTM; supplied by COPAN), stored at 4°C and shipped to central laboratories within 5 days of collection, aliquoted, and stored at -80°C until tested. This trial complied with the Declaration of Helsinki, the International Council on Harmonization Guidelines for Good Clinical Practice, and applicable local regulations. The study protocol was approved by Western IRB (WIRB). Participants were enrolled at ten sites across six states: North Carolina, California, Washington, Texas, Florida, and Louisiana. All study participants provided written informed consent. Doses administered in this study were 200 mg, 400 mg, or 800 mg twice daily (BID). Randomization was 1:1 for the initial 200mg group and 3:1 for both 400 and 800mg groups. A complete list of eligibility criteria is available at clinicaltrials.gov (NCT04405570).

### Biocontainment and Biosafety

All work described here was performed with approved standard operating procedures for SARS-CoV-2 in a biosafety level 3 (BSL-3) facility conforming to requirements recommended in the Microbiological and Biomedical Laboratories, by the U.S. Department of Health and Human Service, the U.S. Public Health Service, and the U.S. Center for Disease Control and Prevention (CDC), and the National Institutes of Health (NIH).

### Measurement of SARS-CoV-2 RNA via Clinical qRT-PCR Assay

To measure viral RNA, a quantitative RT-PCR was performed on NP swab VTM using primers based on the CDC emergency use authorization assay against the N1 region of the nucleocapsid gene (2019-nCoV_N1) at Covance CLS (lower limit of quantification: 1,018 copies/mL)^21^.

### Culture of SARS-CoV-2 Clinical Isolates in Vero Cell Cultures

Infectious SARS-CoV-2 was isolated and cultured in Vero-E6 cells (VERO C1008 [Vero 76, clone E6, Vero E6] (ATCC CRL-1586) expanded and cryopreserved by UNC Tissue Culture Facility using adapted good laboratory practices. Twenty-four hours prior to infection, 24-well culture plates (1.9 cm^2^ area/well) were seeded with 30,000 Vero E6 cells/well/1mL culture medium comprised of Minimal Essential Medium (Gibco), 10% fetal bovine serum (Hyclone), 1X non-essential amino acids (Gibco), 1X antibiotic/antimycotic (Gibco). VTM samples were thawed from −80°C, diluted 1:1 in infection medium (Vero cell culture medium with 4% serum) and added to duplicate Vero cell culture wells (100µl/well). Per batch of samples, quadruplicate negative (medium only) and recombinant SARS-CoV-2 high (multiplicity of infection, MOI = 0.008, 500 plaque forming units, PFU) and low (MOI = 0.0008, 50 PFU) controls were included. After incubation at 37°C for 1hr, input virus was removed, cultures were washed once with 500µl Minimal Essential Medium in order to diminish background RNA associated with the clinical samples not associated with active replication of infectious virus, and 1mL infection medium was added to each well. On 2 and 5 days post-cell culture infection (dpi), 200µl of culture medium was collected from each well and thermally inactivated at 65°C for 35 minutes. Culture positivity was determined by measuring viral RNA in heat-inactivated culture medium by qRT-PCR using the Abbott m2000sp/rt quantitative SARS-CoV-2 RNA assay (limit of quantification: 100 copies/mL) in the UW Retrovirology/Clinical Trials GCLP Lab^22,23^. The absolute limit of culture detection was 50 PFU (MOI 0.0008) using two criteria for positivity: (i) SARS-CoV-2 RNA copies >1,000/mL in supernatant at 2dpi, or (ii) a relative change of >5 in RNA copy number from 2 to 5 dpi (i.e., [copies_5dpi⁄copies_2dpi]>5). The rationale for criteria (i) was to capture culture positivity for samples with abundant virus replication on 2dpi such that increases in copy number would not be observed between 2 and 5 post-infection. For criteria (ii), we aimed to define active replication of infectious virus from clinical samples by measuring an increase in viral RNA from 2 to 5dpi in our assay. In addition, this metric aims to reduce the potential for false positives resulting from lingering contaminating viral RNA that was not removed by washing and is not associated with infectious virus.

### Determining the Presence of Infectious SARS-CoV-2 by Observation of Cytopathic Effect

On 2 and 5dpi, Vero cell culture wells infected with clinical samples or controls were observed for virus-induced cytopathic effect (CPE) by phase contrast microscopy. Briefly, using a 10x objective, each well was scanned for cytopathic effect and the percentage of CPE was estimated and recorded per well. Samples were positive for infectious virus positivity as determined by CPE if: (i) the difference in average CPE between mock and clinical sample wells was greater than 20% or (ii) if CPE increased >10% from 2 to 5dpi. Like the criteria above for RT-PCR-based assays for infectious virus positivity, the CPE-based assay criteria were designed to capture samples that had abundant infectious virus such that maximal CPE was attained by 2dpi as well as samples with lower levels of infectious virus and measurable increases in CPE from 2 to 5dpi in the assay.

### Illumina sequencing of purified viral RNA

Purified RNA from expanded viral cultures was submitted to the Vanderbilt University Medical Center (VUMC) core facility, Vanderbilt Technologies for Advanced Genomics (VANTAGE). For RNA-Seq, total RNA underwent poly(A) selection followed by NovaSeq PE150 sequencing (Illumina) at 15 million reads per sample. Reads were then aligned to the reference genome (MT020881.1) and an in-house pipeline, *CoVariant*, was used to identify, quantify, and annotate mutations. Locations of amino acid changes were confirmed through sequence alignment using MacVector and CLC Workbench (QIAGEN). Sequencing data can be found via Sequence Read Archive (SRA) accession PRJNA1043164.

### SARS-CoV2 ELISA

The site-specifically biotinylated halo-tagged SARS-CoV-2 spike RBD (331-528 aa, P0DTC2.1) was produced as described before ^24,25^. The SARS-CoV-2 N antigen (245-364 aa, P0DTC9.1) with a halo-tag was produced in mammalian Expi293 cells and site-specifically biotinylated as described for RBD antigen. Titration ELISAs were carried out as previously described^26^. In brief, streptavidin was coated at 4 μg/mL in Tris-Buffered Saline (TBS) pH 7.4 for 1 h at 37°C followed by blocking with Non-Animal Protein-BLOCKER (GBiosciences). Biotinylated RBD or N antigen was added at 1 μg/mL at 37°C for 30 min. All plasma samples were heat-inactivated before use to minimize the risk of residual virus in serum and then incubated at serial dilution (1:33 to 8100). The plates were subjected to multiple washes and incubated with horseradish peroxidase-conjugated secondary Goat Anti-Human secondary IgG (109-035-008), IgM (109-035-043), or IgA (109-035-011) Abs from Jackson ImmunoResearch at a dilution of 1:40,000 in 3% milk at 37°C for 1 h. The resulting plate was washed again, and 3,3′,5,5’ - Tetramethylbenzidine (TMB) Liquid Substrate (T0440, Sigma-Aldrich) was added for optical density (OD) measurement at 405 nm after stopping the reaction with 1 N HCl.

### Multiplex surrogate SARS-COV-2 neutralization assay

We measured the ACE2 blocking antibodies against a panel of spike proteins from SARS-CoV-2 reference strain and circulating variants, including (WT), Alpha, Beta, Delta, IHU (B.1.640.2), BA.1, (BA.1+L452R), (BA.1+R346K), (BA.2), and (BA.3) using the multiplexed MSD V-PLEX SARS-CoV-2 Panel 25 (ACE2) as previously described^25^. Briefly, plates were blocked with MSD Blocker A for 30 minutes and washed three times before adding reference standard, controls, and heat-inactivated samples diluted 1:50 in the diluent buffer. After incubating the plates for 1 hour with shaking at 700 rpm, a 0.25μg/ml solution of MSD SULFO-tag conjugated ACE2 was added and incubated for 1 hour with shaking. Finally, the plates were washed and read with a MESO QuickPlex SQ 120 instrument. The ACE2 blocking activity was reported as a percentage of inhibition calculated based on the equation ((1 – Average Sample ECL Signal / Average ECL signal of blank well) x 100).

### Statistical Analyses

We used Wilcoxon signed-rank tests to compare day 1 SARS-CoV-2 RNA levels (log_10_ copies/mL) between swab types (Figure 1B). Separately for each swab type, SARS-CoV-2 RNA levels (log_10_ copies/mL) were compared by infectious virus status (positive/negative) combined over days 1, 3, and 5 using a Wilcoxon rank-sum test for clustered data (Datta-Satten method) to account for repeated measures across the 3 days (Figure 1C)^27^. The number of changes on day 3 for synonymous, transition, and transversion mutations were compared between treatment groups (MOV and Placebo) using a Wilcoxon rank-sum test (Figure 2B)^28^. Jonckheere-Terpstra (JT) tests for trend were used to evaluate Molnupiravir dose-response trends (800mg, 400mg, Placebo) in antibody titer levels at day 7 and day 28 separately for each antibody type and time point (Figure 3), and to evaluate the dose-level trend in ACE2 antibody blocking levels on day 28 for the Wuhan strain for participants who were seronegative by RBD IgG on day 1 (Figure 4). Missing data were excluded, and p-values are presented with no adjustment for multiplicity. Analyses were conducted in Windows SAS version 9.4 (Cary, NC) and Windows R versions 4.0.4 or 4.1.2.

## Supplementary Figure Legends

**Supplementary Figure 1.**
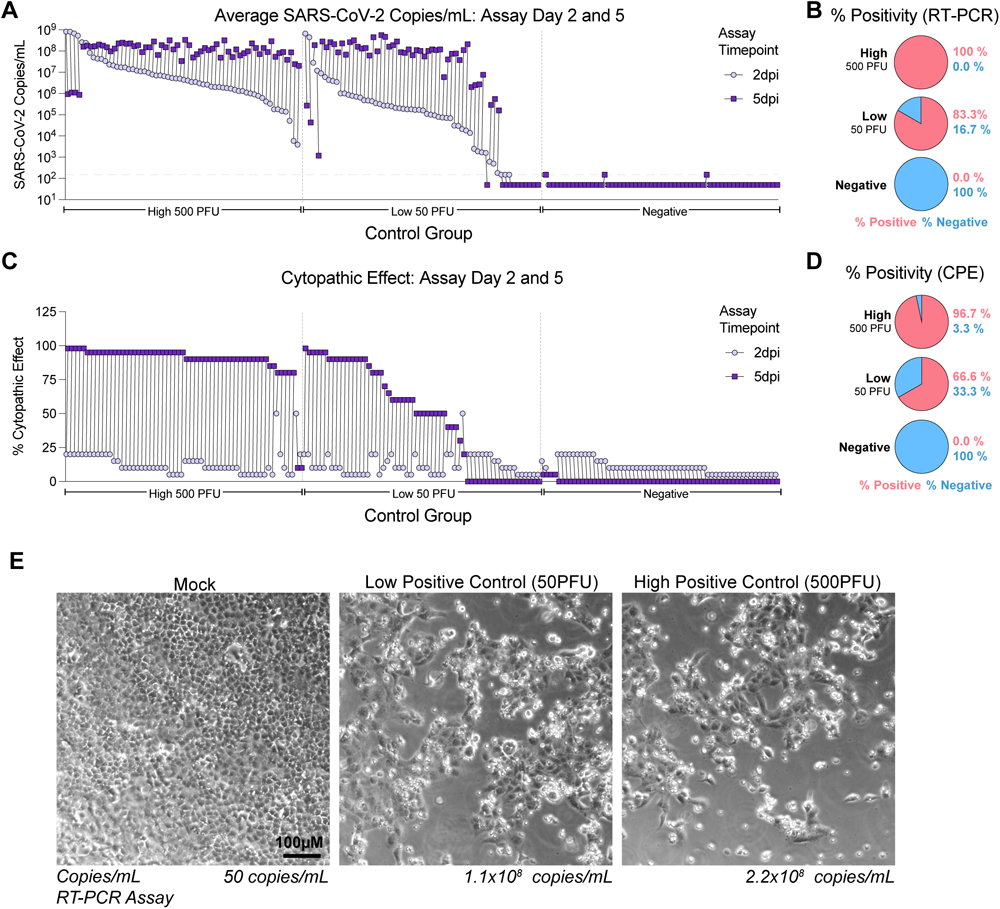
Infectivity data by Culture PCR and cytopathic effect. (**CPE) for recombinant SARS-CoV-2 controls. (A)** Culture PCR data for High, Low and Negative SARS-CoV-2 controls. **(B)** Proportion of Culture PCR positive samples for High, Low and Negative SARS-CoV-2 controls. **(C)** CPE data for High, Low and Negative SARS-CoV-2 controls. **(D)** Proportion of CPE positive samples for High, Low and Negative SARS-CoV-2 controls. **(E)** Photomicrographs of example CPE from High, Low and Negative SARS-CoV-2 controls.

**Supplementary Figure 2.**
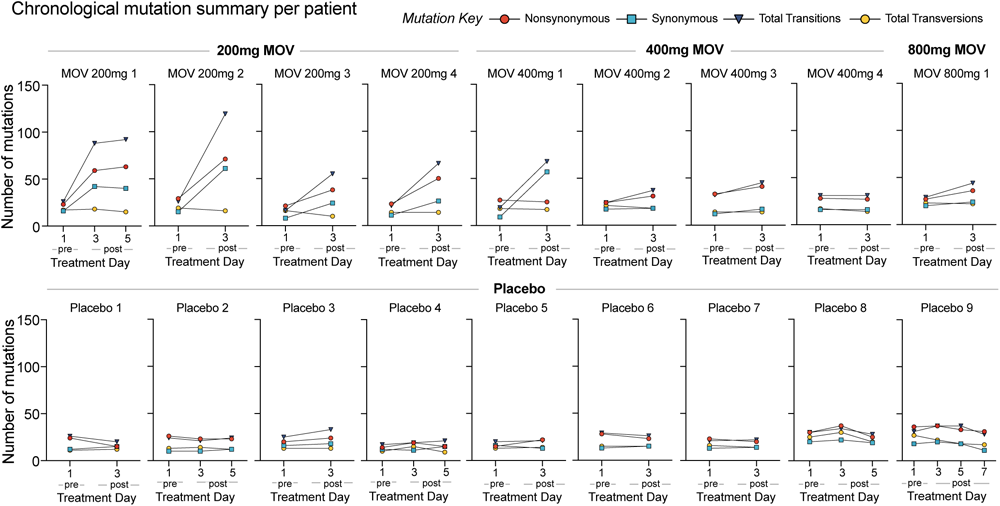
Number and identity of mutations in infectious virus populations isolated from MOV and Placebo treated participants. The number of nonsynonymous, synonymous, transition and transversion mutations are shown per participant treated with 200, 400, 800mg MOV or Placebo.

**Supplementary Figure 3.**
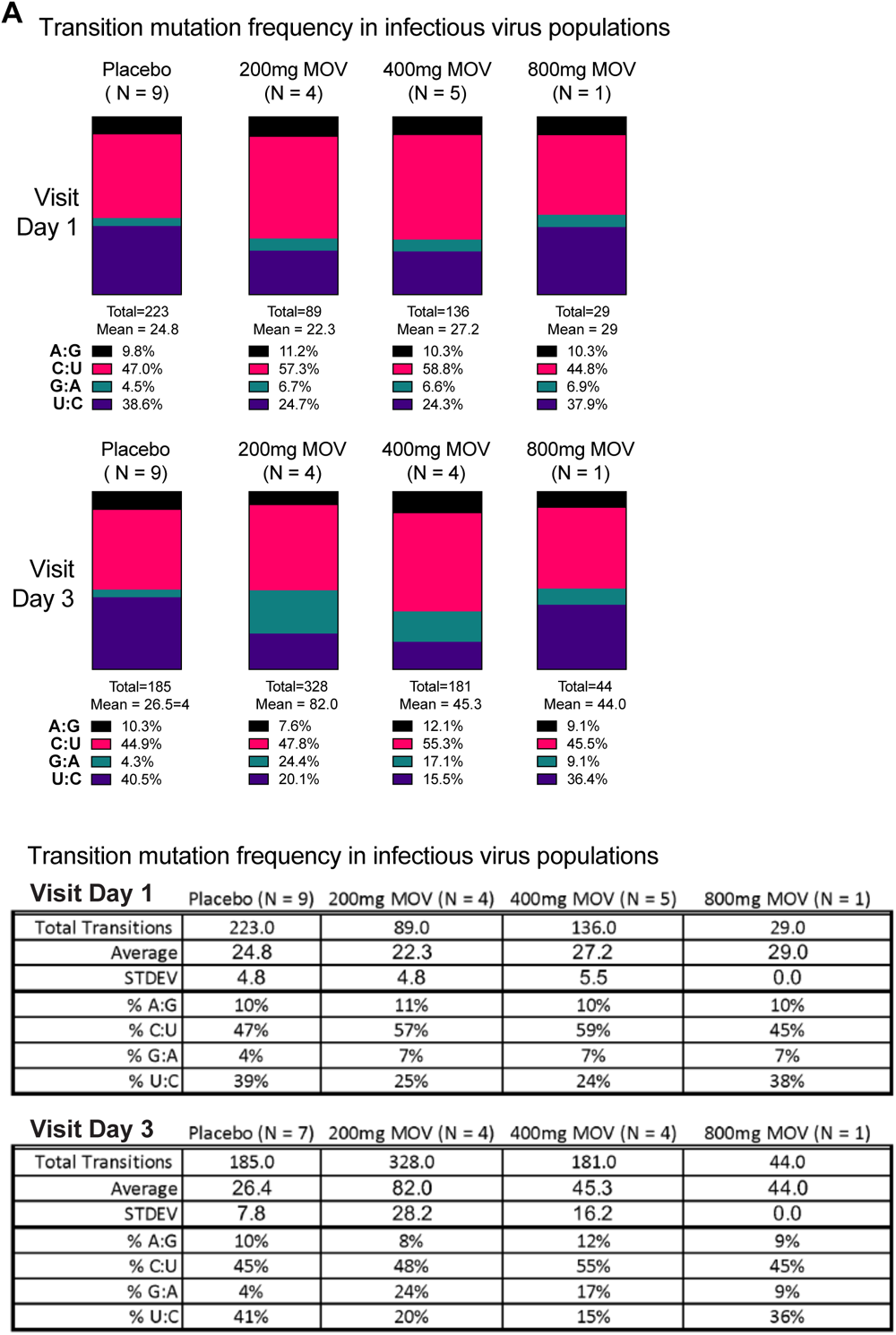
Transition mutation frequency in infectious virus populations from MOV and Placebo treated participants. (A) Frequency of A:G, C:U, G:A and T:C transitions per study group.

**Supplementary Figure 4.**
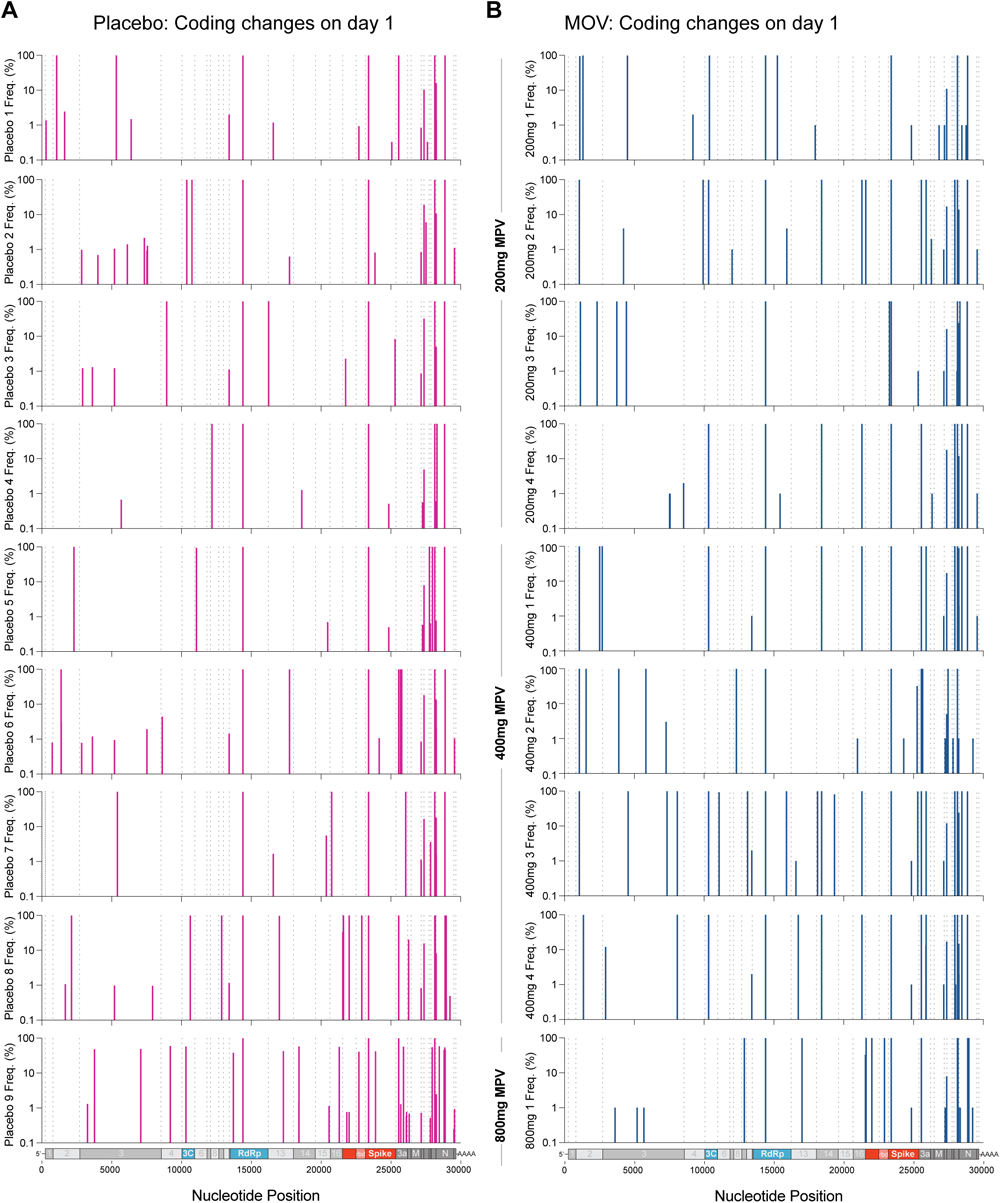
The location and frequency of coding changes on visit day 1 in infectious virus populations isolated from Placebo and MOV-treated participants. (A) The location and frequency of amino acid coding changes among infectious virus populations isolated from Placebo treated (A) or MOV-treated (B) participants on visit Day 1.

**Supplementary Figure 5.**
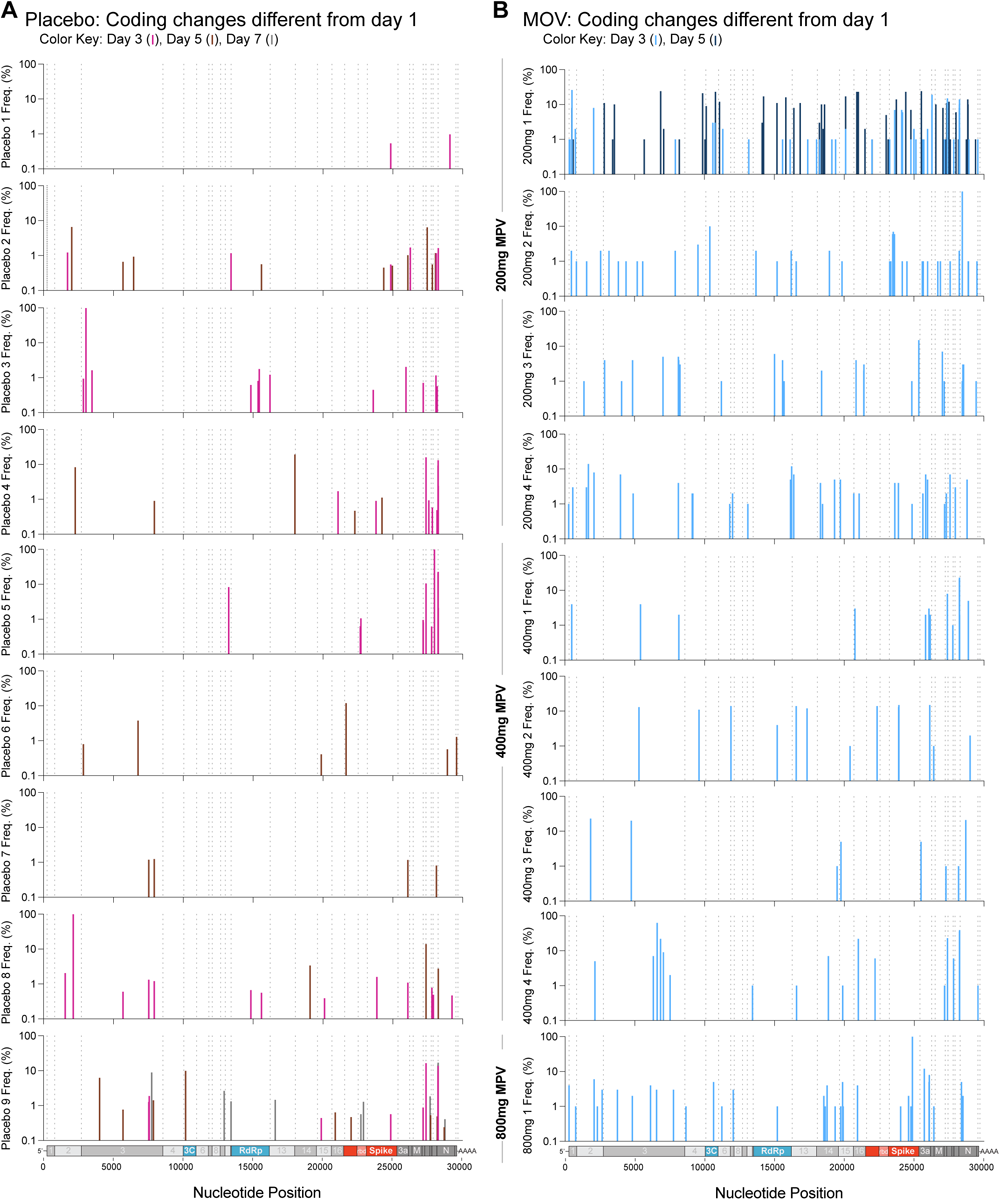
The location and frequency of coding changes different than day 1 in infectious virus populations isolated from Placebo and MOV-treated participants on visit days 3, 5, and 7. (A) The location and frequency of amino acid coding changes among infectious virus populations isolated from Placebo treated (A) or MOV-treated (B) participants on visit Day 3, 5 and 7.

**Supplementary Figure 6.**
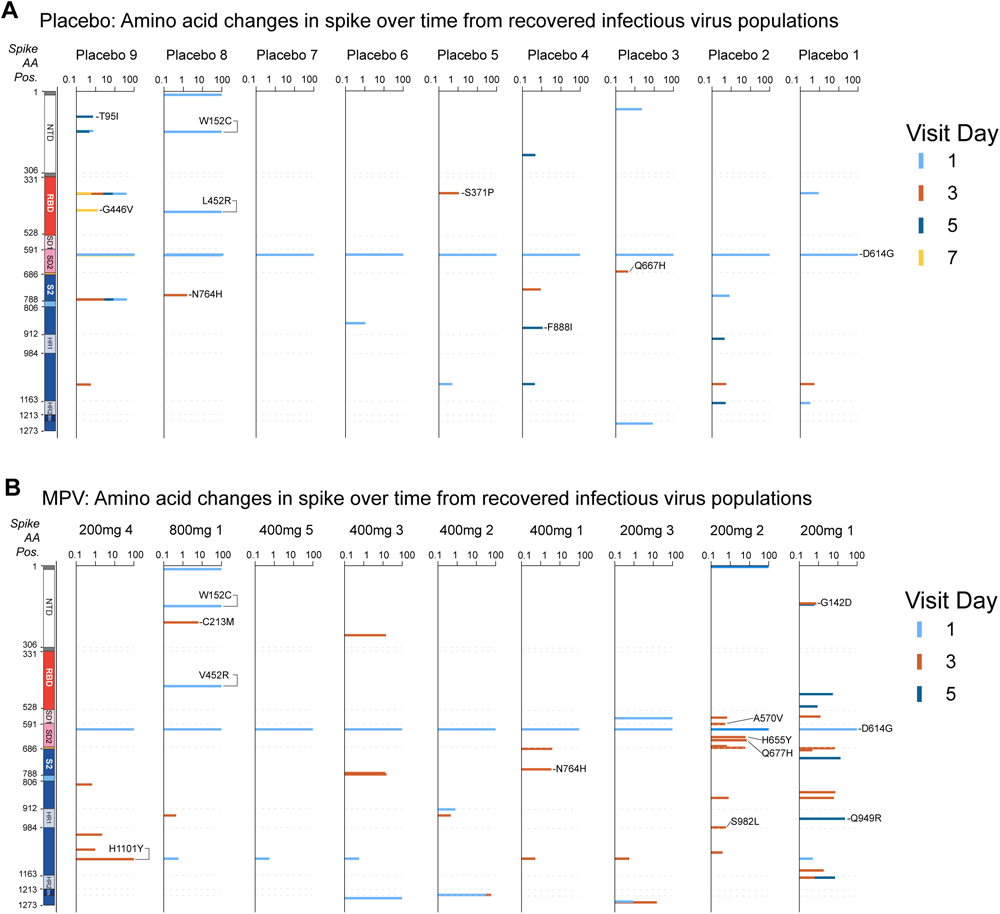
Amino acid changes in spike over time. Amino acid changes in the SARS-CoV-2 spike protein in infectious virus isolated from Placebo **(A)** and MOV **(B)** treated participants.

